# What do we know about violence against women in pandemic times? Insights based on search trends

**DOI:** 10.1101/2021.05.26.21257747

**Authors:** Rafaela Ferreira Guatimosim, Ana Luiza Silva Teles, Fabiano Franca Loureiro, Antônio Geraldo da Silva, Débora Marques de Miranda, Leandro Fernandes Malloy-Diniz

## Abstract

**Purpose:** This short communication aims to assess the situation of domestic violence against women in Brazil during social isolation due to the COVID-19 pandemic.

**Methods:** We extracted data from Google Trends showing the magnitude of searches on the topics domestic violence and complaint and then compared with the data of the complaint reports.

**Results:** Searches on Google containing those terms have increased while the complaints reports against domestic violence have decreased.

**Conclusion:** The growth of searches about domestic violence and domestic violence complaints indicates the possibility of a real rise in this type of violence in Brazil.

## Introduction

Psychological Distress has been considered among the most important outcomes of COVID-19 pandemics (Silva et al., 2020). As pointed by Connor et al. (2020) gender disparities in risks and health implications are likely to be amplified during the Covid-19 pandemic. Women, since the beginning of the COVID-19 pandemic, have been associated with a greater psychological impact and higher levels of stress, anxiety, and depression, as shown in the study by Wang (2020). There are several factors that may increase the impact of pandemics in women’s mental health, which includes gender inequalities strengthening work overloads at home and gender violence (Souza et al., 2021). In a recent systematic review, Piquero et al. (2021) analyzed 37 studies and found that at least 29 reported a statistically significant association between the stay at home police and the increase in domestic violence. In this context, the United Nations Association (UN) has warned about this problem, highlighting the need to promote protection strategies for women (Guterres, 2020 & United Nations Population Found, 2020).

This situation is alarming since victims of domestic violence are often caught in unmanageable personal situations that make it almost impossible to take effective legal action (Artz, 2011). In the study conducted by Artz (2011) in which she interviewed 503 women, it was shown that fear of the perpetrator and coercion to withdraw legal actions were significant factors affecting women’s decisions to return to court. Artz (2011) also reports that of the thousands of women who begin the process of obtaining protection orders in the U.S., less than half return to court to obtain final requests.

In order to better examine the Brazilian scenario regarding the situation of women, Google Trends was used, a free access tool, which has already been used by several scientific researches (Nuti, 2014; Tsao, 2020 & Vosen, 2011). In the study by Vosen (2011) Google Trends in almost all experiments conducted on consumption-related research proved to be better than conventional survey-based indicators. In Tsao’s (2020) systemic-review, It is shown that Google Trends searches from two weeks earlier can be used to model the number of COVID-19 cases. There is evidence that the tool is useful for addressing health-related phenomena and is even acknowledged as promising by the Institute of Medicine in ways that complement current research and findings (Nuti, 2014). In addition, the data enabled by Trends provide relevant information on the behavior of the population, since research trends can expose and help to predict social phenomena (Nuti, 2014 & Vosen, 2011). This predictive aspect is also useful since the verified behaviors may reappear in the future under similar conditions or contingencies. Such scenario highlights the urgency of this communication, about domestic violence against women, and serves as an incentive for public policies to be created and/or strengthened in order to elaborate preventive and care mechanisms for the victims.

Our study aims to further investigate research trends in order to better examine and address domestic violence in a pandemic context

## Material and methods

On February 28, 2021, we gathered the data released by Google Trends in celebration of International Women’s Day. We chose to search portuguese terms related to women violence topics. We downloaded the data for the following search input: [how to report + domestic violence] and [domestic violence]. The data collected in Brazil were from 2004 to 2021 without specifying the query category.

Google Trends is a tool that provides search interest figures relative to its highest point. The value of 100 represents the peak popularity of the term. This means that the data is indexed so that 100 represents the greatest research interest, and the additional scores are made proportionally in relation to the maximum point. We describe the search interest frequencies yearly across time, and then compared the search trends data with the reports on crimes against women by FBSP (2020).

## Results

In Brazil, in 2020 the number of searches for [how to report + domestic violence] reached its highest point in the last 17 years, as did the search interest for “domestic violence,” which also reached its highest point in the last 11 years (Google Trends, 2021).

The peak of the search interest of reporting domestic violence was reached in March 2020, the first month of extensive social distancing measures in most states in Brazil. Also noteworthy is the result from May of the same year when 98 was reached. The highest point of search interest for ‘domestic violence’ was reached in July 2020, when 28 was registered.

Graph 1 displays data from Google Trends regarding search interest for the terms ‘Domestic Violence’ and ‘how to report + domestic violence’ in Brazil from 03/01/2015 to 05/04/2021. The dotted line marks the month of February, the month in which the beginning of the Public Health Emergency of National Importance (ESPIN) was demarcated in the country as a result of COVID-19 that was declared through Ordinance No. 188/ GM/MS.

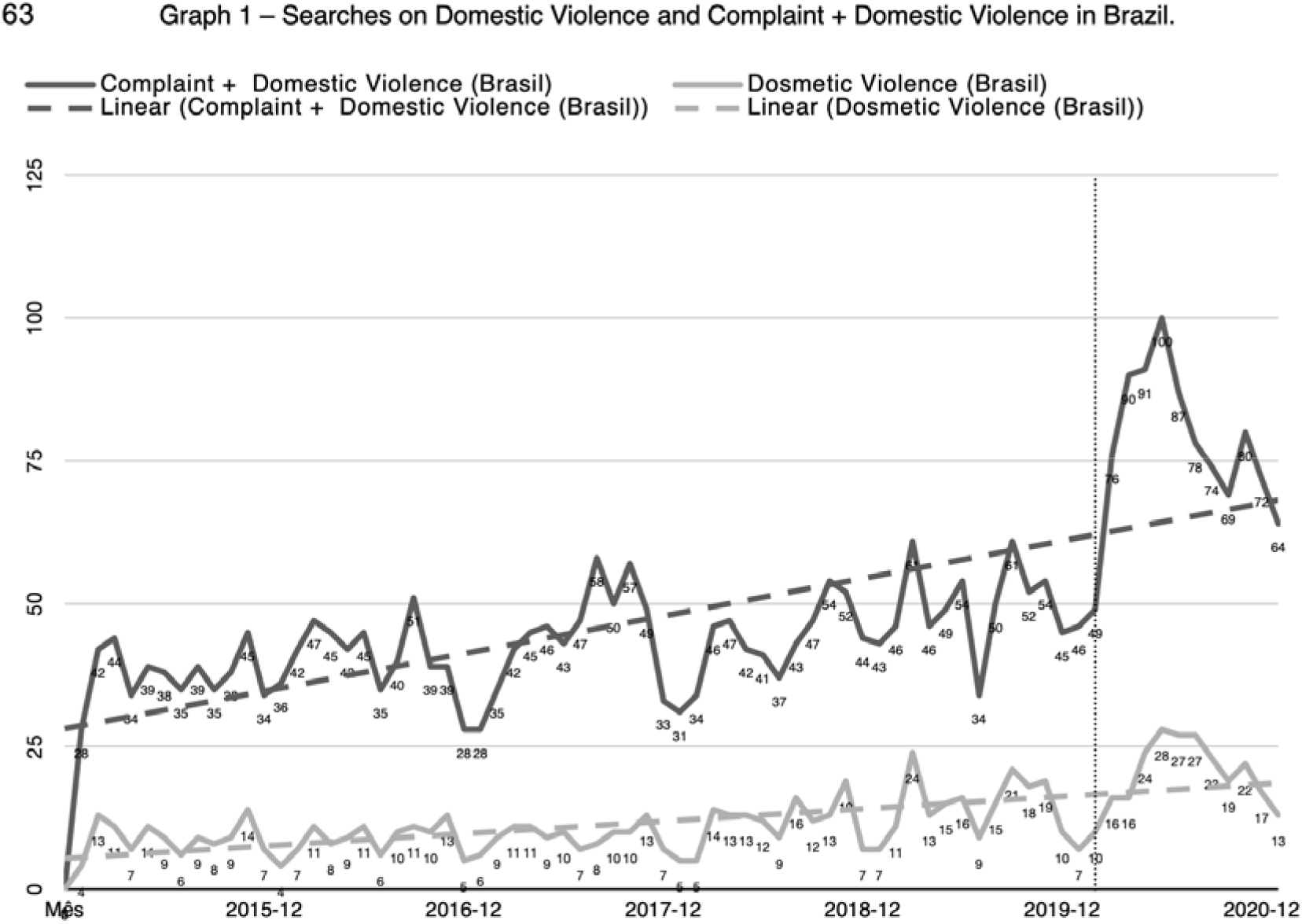

## Discussion

These data show a greater interest in “*how to denounce*’’ than in the search of “*what domestic violence is*”. It also reveals an increase in searches about domestic violence and contrasts with the data collected by the Brazilian Forum for Public Safety (FBSP), which states a reduction in a number of crimes against women (Fórum Brasileiro de Segurança Pública, 2020). According to the FBSP, the crime of bodily injury had a reduction in almost all Brazilian states, showing a drop of 9.9% when comparing the first semester of 2020 to that from 2019. There was also a reduction in threatening crimes by 15.8%. The records of rape fell by 23.8% in the same period. Nevertheless, in contrast to the other data presented, feminicide had an increase of 1.9% in the same period. Another data that draws attention is the 3.8% increase in the total number of calls to 190 recorded under the nature of domestic violence in the first semester when compared to that of 2019.

To this we must add that, as stated by Artz (2011) reporting domestic violence may have a ‘boomerang effect’, in which seeking help amplifies threatening behaviors by the aggressor. Artz (2011) explains that unlike most types of crime, the perpetrator is motivated to retaliate when the victim seeks legal help. Nevertheless, victims need to be made aware that the intensity and brutality of domestic violence increases with time, and psychological and mental abuse escalates into physical violence (Artz, 2011).

Given this complex picture, in which the search for denunciations is increasing and the records of violence are decreasing, the need for attention to the situation in which Brazilian women find themselves becomes evident. Although the social isolation measures are extremely important and necessary for the management of the pandemic, the recommendations to stay at home have reinforced the coexistence with potential aggressors, as have been reported by several studies (Piquero et al., 2021). In addition to limiting their access to protection networks: travel restrictions have prevented and/or made it difficult for the victims to leave the violent environment, as well as in many cases preventing them from having access to family members, shelters, temporary shelter homes, and hotels. Moreover, in the face of the current stressful period, there is a tendency for family tensions to worsen, especially in those with a history of recurring domestic violence, which become even more prone to such situations (Viero, 2021).

We also highlight an important fact: in 2020, the unemployment rate in the country increased substantially, and the unemployment rate among women was 16.4% in the last quarter of the year, according to data from the IBGE (Barros, 2021), which ends up reflecting on other problems, such as economic dependence and, with this, a greater vulnerability to the aggressor. Furthermore, during the pandemic, many women ended up being overburdened due to the accumulation of duties, both among women with paid jobs and among those without, who also end up accumulating more domestic workload and caring for other people such as children and family members (Power, 2020).

According to Dang and Nguyen (2021) Covid-19 pandemics increased the gender’ gap in the workforce since many women became unemployed and expected income reduction in comparison to men. Women in financial vulnerability are more likely to be victims of partner violence (Pereira and Gaspar, 2021) and we can hypothesize that in those scenarios of financial dependence, reporting domestic violence to authorities becomes less probable.

One has a scenario that feminicide (a crime that is harder to hide its occurrence) has increased, as have calls to police and research regarding how to report it, while the records of various crimes associated with domestic violence have decreased (Fórum Brasileiro de Segurança Pública, 2020). In addition, there is also the fact that a global increase in domestic violence crimes associated with the pandemic has been reported by the UN (Guterres, 2020 & United Nations Population Found, 2020). From this setting, it is speculated that social isolation contributes to the underreporting of statistics, and that women are facing greater difficulties in making complaints and accessing specialized women’s services. This hypothesis highlights the need for more research on the current context in which Brazilian women find themselves, and for more support to make it possible to file complaints.

This study presents some important limitations. Data from different sources were cross-referenced, which can already lead to biases or inaccuracies in the analysis. We also recognize that these statistics involve different variables, and to attribute a behavior or effect to only one of these variables is also leading to inaccuracies. While acknowledging the possible flaws, it is still pointed out that the hypothesis raised from the data is alarming. The purpose of this brief communication is to draw attention to this hypothesis, which by the mere fact that it is possible, calls for further studies.

One of the strengths of the study consists of the fact there is indirect data that might reflect an approach less susceptible to under notification. Therefore, women’s protection services must be redirected to actions that can be effective even remotely, such as campaigns that highlight the existence, importance, and effectiveness of the channels for denunciation: the number 180 and 100, stimulating the woman to denounce, as well as people close to her who see what is happening. It is also possible to report violence through the “Proteja Brasil” app, developed by the United Nations Children’s Fund (Unicef) and by the then Secretariat of Human Rights of the Presidency of the Republic (now the Ministry of Human Rights), which allows not only making the complaint directly through the application, but also informing the location of the protection agencies in the main capitals, besides also containing information about the different violations. Additionally, based on the information and numbers found in Google Trends, it is also important to disseminate the behaviors that characterize domestic violence for the greater knowledge of the population, since, except for physical and sexual violence, the others can manifest themselves in very subtle ways since they do not leave “marks”.

## Data Availability

All data is available on Google Trends. The instructions on how to get the data are presented in the article.

https://trends.google.com.br/trends/story/BR_cu_Jmj4q3ABAACuQM_en.

## Declarations

This work was supported Panamerican Health Organization (PAHO) [grant number SCON2020-00202] and CNPQ [grant number 401542/2020-3] in task force with Brazilian Association of Psychiatry (ABP), Brazilian Association of Impulsivity and Dual Patology (ABIPD) and SAMBE Research Group.

## Conflicts of interest/ Competing interests

None.

## Notes

### Competing Interest Statement

The authors have declared no competing interest.

